# Women’s Sociocultural Representations of HIV Pre-Exposure Prophylaxis and Intimate Partner Violence in Heterosexual Relationships: A Qualitative Story Method

**DOI:** 10.1101/2025.02.20.25322597

**Authors:** Tiara C. Willie, Laurel Sharpless, Deja Knight, Aashna Shah, Amina Antar, Z. Thomasina Watts, Kamila A. Alexander, Trace Kershaw

**Author notes:** **Corresponding author:** Tiara C. Willie, PhD, MA; Department of Mental Health, Johns Hopkins Bloomberg School of Public Health, Baltimore, MD, USA.

## Abstract

**Objective:** The current study sought to assess women’s understanding of pre-exposure prophylaxis interest and initiation in heterosexually-active relationships with and without histories of intimate partner violence.

**Design:** 2017-18 prospective cohort study with an embedded, story completion exercise in the 90-day follow-up such that participants were randomized to receive one of two versions of a story stem based on whether they experienced intimate partner violence.

**Setting:** Connecticut.

**Participants:** 132 heterosexually-active, cisgender adult women residing in the state of Connecticut.

**Primary outcomes measures:** Our primary outcome included identifying principal narratives that describes women’s conceptualizations of pre-exposure prophylaxis interest and initiation in heterosexually-active relationships.

**Results:** Using both story mapping and thematic analysis techniques, four principal narratives were identified across the stories: 1) the *Informed and Empowered PrEP User*, 2) the *Clandestine PrEP User*, 3) the *Hesitant PrEP Contemplator*, and 4) the *Disenfranchised PrEP Non-User*. These novel narratives provide insights on how social, clinical, and interpersonal factors are underpinning heterosexually-active, cisgender adult women’s ability to display interest and initiate pre-exposure prophylaxis in their relationships.

**Conclusions:** If our findings are replicated in studies in different settings, it will provide substantial support for future prevention interventions adopting empowerment-centered approaches to refocus women’s needs in the context of PrEP initiation and healthy relationships.

## INTRODUCTION

The inequitable implementation of HIV pre-exposure prophylaxis (PrEP) in the United States places marginalized women at greater risk for acquiring HIV through heterosexual contact^1–3^ and women experiencing intimate partner violence (IPV).^4–7^ To date, PrEP coverage is lowest among females than males.^8,9^ Although limited research has examined PrEP coverage among women experiencing IPV, women experiencing IPV are often forced to choose relationship safety over HIV prevention due to the sociocultural consequences of IPV.^10,11^ Multilevel factors are contributing to disparities in PrEP coverage among women,^12–14^ thus it is important to identify modifiable factors to improve PrEP uptake and initiation.

While structural determinants are critical to address disparities in PrEP use,^4,15,16^ individual determinants may also weaken women’s motivations and interest in PrEP. For example, women with strong skepticism of PrEP feasibility in heterosexually active relationships may have low interest in PrEP.^2^ However, most research regarding women’s interest in PrEP is limited. Specifically, research examining barriers to PrEP interest among women primarily examine individual-level barriers such as low HIV perceived risk and limited HIV knowledge.^13,17^ While low perceived risk and limited HIV knowledge have been associated with low PrEP intentions among women,^18,19^ prior research on HIV prevention among women indicate that belief systems and understandings can also result in low engagement in HIV prevention behaviors.^20,21^ Specifically, studies indicated that women were less likely to use male condoms if they believed that condoms: would negatively impact sex,^22,23^ were embarrassing,^24,25^ and did not work well.^26^ Extending prior research, it is possible that women’s belief systems, conceptualizations, and understandings of PrEP in heterosexually active relationships could shape their interest in PrEP and be an important prevention effort for women.

Qualitative story completion presents a unique opportunity to examine women’s social and cultural understandings of PrEP use in heterosexually active relationships. Qualitative story completion is a research method in which participants are provided with a story stem and are asked to write a response.^27–29^ Primarily used in psychology,^29^ qualitative story completions are beneficial to receive participants’ responses on socially undesirable and sensitive topics in a safe way such as infidelity, sexual aggression, and sexual experimentation.^29,30^ Therefore, the current study aims to assess women’s understanding of PrEP in heterosexually active relationships with and without histories of IPV.

## METHODS

### Procedures

Two hundred eighteen women were recruited from August 2017 to April 2018 to participate in a prospective cohort study exploring factors influencing engagement in the PrEP care continuum among women with and without IPV experiences. These methods are described in prior publications.^5,6^ Flyers were posted online on Craigslist and Facebook, and throughout the community in beauty salons and community health clinics. Participants had the option to be screened for eligibility either through online survey or over the phone. Women were eligible for the study based on the following criteria: (1) aged 18 to 35 years, (2) reported at least one of the sexual risk indicators for PrEP according to the CDC clinical summary guidelines^31^ (i.e., in the past 6 months had unprotected sex with a male partner, HIV-positive sexual partner, recent STI, 2 or more sexual partners, or transactional sex), (3) spoke English and/ or Spanish, and (4) resided in Connecticut. This study also oversampled women experiencing IPV to gain a better understanding of how recent IPV impacted women’s sexual health. Women were considered experiencing IPV if they reported at least one physical and/or sexual abuse experience with a male partner in the past 6 months. Self-reported HIV status was ascertained during the baseline survey, of which all the women reported an HIV negative status.

Women provided written informed consent and were asked to complete a baseline and 90-day follow-up survey administered through Qualtrics either in-person at the research office or online. In each survey, participants were provided patient education information about PrEP. At the end of each survey, participants were compensated $25 and provided a list of community resources (e.g., domestic violence agencies). The [institution masked for peer review] IRB approved all study procedures.

#### Quantitative Measures

**Sociodemographics**. During the baseline survey, participants self-reported the following sociodemographic characteristics: age (in years); sexual identity (heterosexual, bisexual, lesbian, or queer), employment (full-time, part-time, unemployed, not in workforce), and race and ethnicity (Non-Hispanic Black or African American, Non-Hispanic White, Hispanic, Non-Hispanic American Indian/Alaska Native, Native Hawaiian or Other Pacific Islander, Asian, Multiracial, and Another Racial Group).

**Intimate Partner Violence**. Physical and sexual IPV within the past 6 months was assessed using two separate scales. The 12-item physical assault subscale of the Conflict Tactics Scale-2 (CTS-2) was used to assess minor and severe forms of physical IPV experiences.^32^ Responses were recoded according to Straus et al. (2003) (i.e., Never = 0; Once = 1; Twice = 2; 3–5 times = 4; 6–10 times = 8; 10–20 times = 15; >20 times = 25).^33^ Example item is “Threw something at you that could hurt.” The Cronbach’s alpha was 0.90. The short form of the Sexual Experiences Survey (SES) was used to assess sexual IPV experiences.^34^ Example item is “Has your partner kissed or touched you sexually or removed some of your clothes when you did not want to?” Participants were asked to report the frequency with options ranging from 0 to 3+. The Cronbach’s alpha was 0.88. Responses to each scale was summed and then dichotomized to represent (0=No to IPV experience, 1=Yes to IPV experience).

#### Story Completion Exercise

The 90-day follow-up survey contained the story completion exercise. Using 1:1 randomization based on their experience of IPV, participants were assigned to one of two versions of a third person story designed to suggest women’s perspectives of HIV prevention in relationships with IPV experiences. Randomizing based on IPV experience allows an equal number of participants to receive each version of the story and reduces potential selection bias and creates more comparable groups (**Figure 1**). In Version A, the female character attends a doctor’s appointment after suspecting that the male character is committing infidelity. In Version B was identical to Version A, except the male character has used physical violence towards the female character in the past.

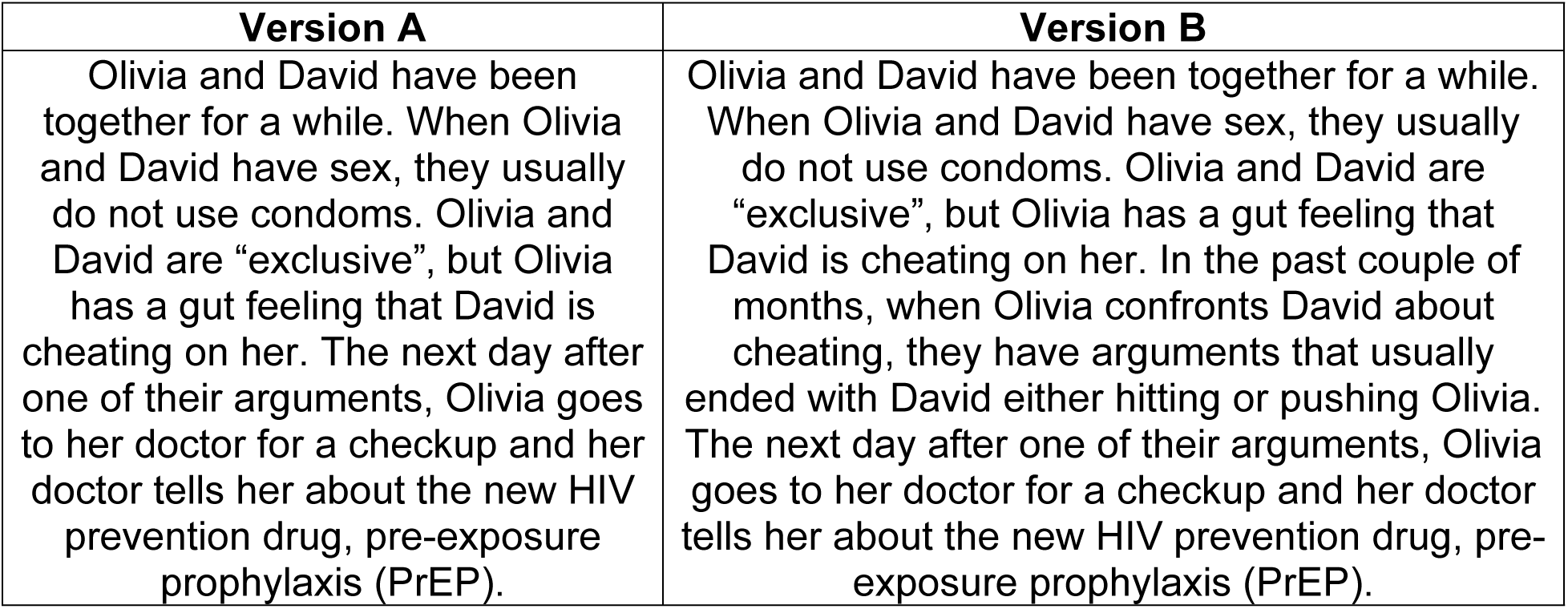

| Version A | Version B |
| --- | --- |
| Olivia and David have been together for a while. When Olivia and David have sex, they usually do not use condoms. Olivia and David are “exclusive”, but Olivia has a gut feeling that David is cheating on her. The next day after one of their arguments, Olivia goes to her doctor for a checkup and her doctor tells her about the new HIV prevention drug, pre-exposure prophylaxis (PrEP). | Olivia and David have been together for a while. When Olivia and David have sex, they usually do not use condoms. Olivia and David are “exclusive”, but Olivia has a gut feeling that David is cheating on her. In the past couple of months, when Olivia confronts David about cheating, they have arguments that usually ended with David either hitting or pushing Olivia. The next day after one of their arguments, Olivia goes to her doctor for a checkup and her doctor tells her about the new HIV prevention drug, pre-exposure prophylaxis (PrEP). |

**Figure 1.**
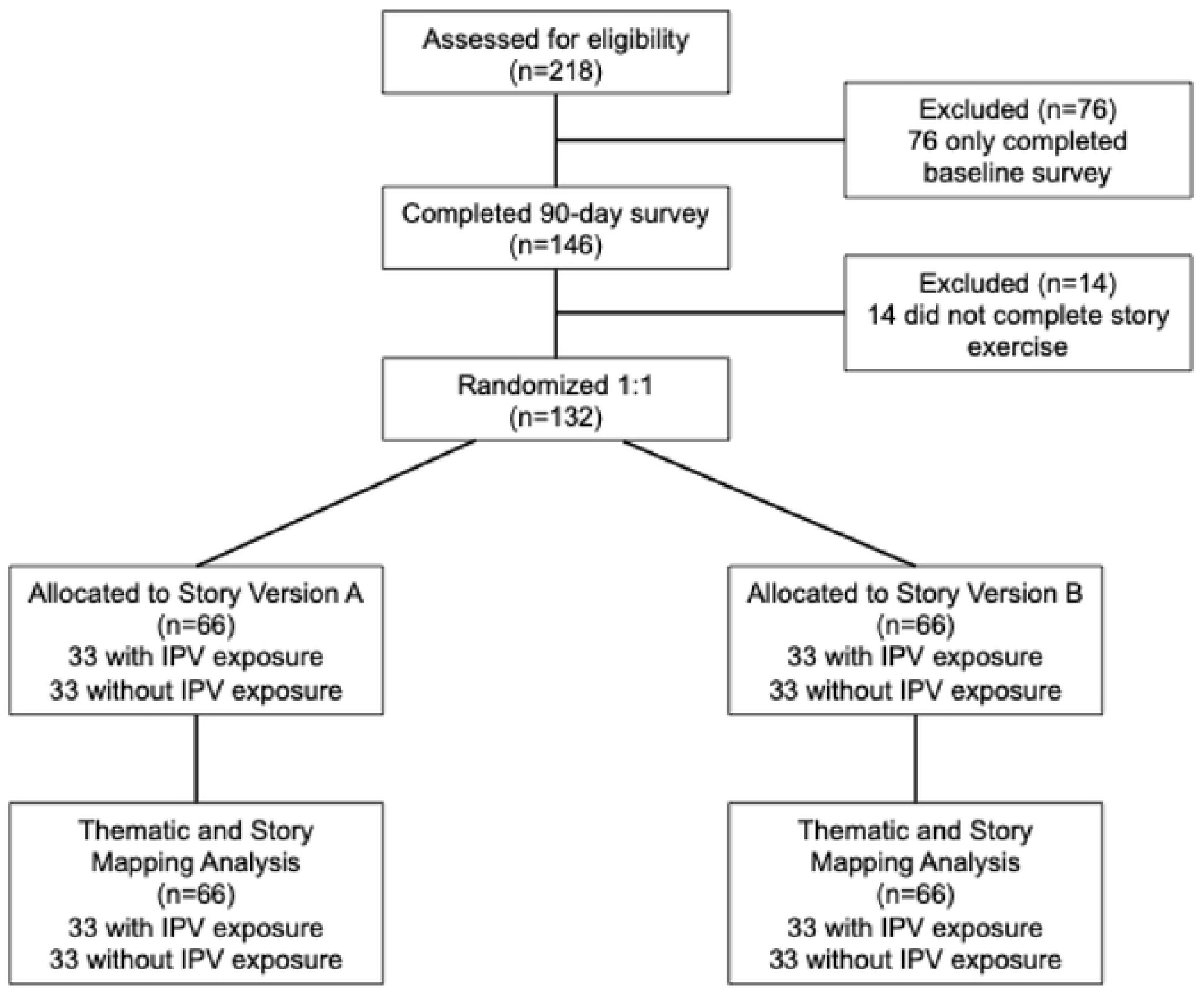
Flow Diagram of Randomized Story Completion Experiment of Two groups Women With and Without Experiences of Intimate Partner Violence, United States, 2017-2018

Each stem was followed by the question “What happens next?” and participants were asked to spend 5 minutes writing the rest of the story.

### Data Analysis

#### Quantitative Data Analysis

During the analysis phase, the research team calculated descriptive statistics (e.g., means, counts, frequencies) among sociodemographic and relationship variables, and qualitative codes. Chi-square tests were used to assess bivariate associations between IPV experience status and qualitative code application. *P*-values<.05 were used to assess significance. The research team conducted all analyses in 2024 by using SPSS version 29^35^ and Dedoose 5.5.^36^

#### Qualitative Data Analysis

The story completion data were analyzed in two different ways: 1) thematic analysis, and 2) story mapping. Using a positivist version of thematic analysis, the stories were coded and analyzed into common themes.^29,30^ With thematic analysis, this approach examined depictions of (a) PrEP as an HIV prevention method, (b) PrEP initiation process, and (c) healthcare providers and male partners influence on women’s PrEP deliberation. Using story mapping, the stories were also coded and analyzed based on how the story unfolded which distinguishes patterns in the actual story elements (i.e., conflict, climax, resolution).^29,30^ With story mapping, this approach identified which common themes occurred in a structured and patterned way in the story.^29,30^

For both analytic approaches, open coding was conducted on 10% of the stories (n=13 stories) by an interdisciplinary team of six coders. The demographics, relationship characteristics, and stem version for each participant were masked from the coders during the coding process. During the open and axial coding stages, codes were discussed, aggregated and condensed in an iterative process among the research team to develop a codebook. The codebook was modified and finalized based on these discussions. All the transcripts were coded based on the finalized codebook by six coders. During consensus team meetings, there were discussions regarding inconsistencies in code application and interpretation and confirming and disconfirming cases. Qualitative data was coded using Dedoose 5.5.^36^

In this study, we used four criteria of trustworthiness for our qualitative analysis: credibility, transferability, dependability, and confirmability.^37,38^ To increase credibility and dependability, we had prolonged engagement in the data (i.e., multiple stories, intensive coding), and managed data triangulation (i.e., stories and surveys assessing similar constructs). To enhance transferability, the research team used purposive sampling to focus on key informants. To enhance confirmability, the Principal Investigators provided a thorough description of the research process.

## RESULTS

### Story Completion Participants

Among the 146 participants who completed their 90-day follow-up survey, 132 stories were able to be analyzed for this current study (66 participants completed either Version A or B). Of the 134 participants, nearly three-fourths of participants were currently employed (**Table 1**). Approximately, 78% of participants identified as heterosexual, and 22% identified as lesbian, bisexual, or queer. More than a third of the participants self-identified themselves as non-Hispanic white (37.1%), followed by non-Hispanic Black or African American (26.5%), Hispanic (15.1%), Multiracial (12.8%), Asian (7.4%), another racial group (1.5%), non-Hispanic American Indian/Alaska Native (0.7%), and Native Hawaiian or Other Pacific Islander (0.7%). Half of the participants (50%) reported experiencing physical, psychological, and sexual IPV in their lifetime.

**Table 1.** Characteristics of 132 Women With and Without Experiences of Intimate Partner Violence Who Participated in the Prospective Cohort and Story Completion Exercise, United States, 2017-2018.

|  |  |
| --- | --- |
| Demographics | N (%) |
| **Age, *M*, *SD*** | 25.5 (5.1) |
| **Employment** |  |
| Employed | 99 (75.0) |
| Unemployed | 33 (25.0) |
| **Sexual Identity** |  |
| Heterosexual | 103 (78.0) |
| Lesbian, Bisexual, or Queer | 29 (22.0) |
| **Race and Ethnicity** |  |
| Non-Hispanic Black or African American | 35 (26.5) |
| Non-Hispanic White | 49 (37.1) |
| Hispanic | 20 (15.1) |
| Non-Hispanic American Indian/Alaska Native | 1 (0.7) |
| Native Hawaiian or Other Pacific Islander | 1 (0.7) |
| Asian | 10 (7.4) |
| Multiracial | 17 (12.8) |
| Another Racial Group | 2 (1.5) |
| **Intimate Partner Violence** |  |
| Yes | 66 (50.0) |
| No | 66 (50.0) |

### Frequencies of Code Application

**Table 2** displays the count and frequency by IPV experience status of the nine commonly used codes to develop the narratives. PrEP decision-making was the most prevalent code with 80 applications, followed by Provider outreach (57 counts), PrEP initiation (53 counts), Partner conversations (32 counts), and IPV (31 counts). Of the five most applied codes, four of these codes tended to be identified in stories written by participants who did not experience IPV than participants with these experiences. The IPV code was most identified by women who had experienced IPV.

**Table 2.**
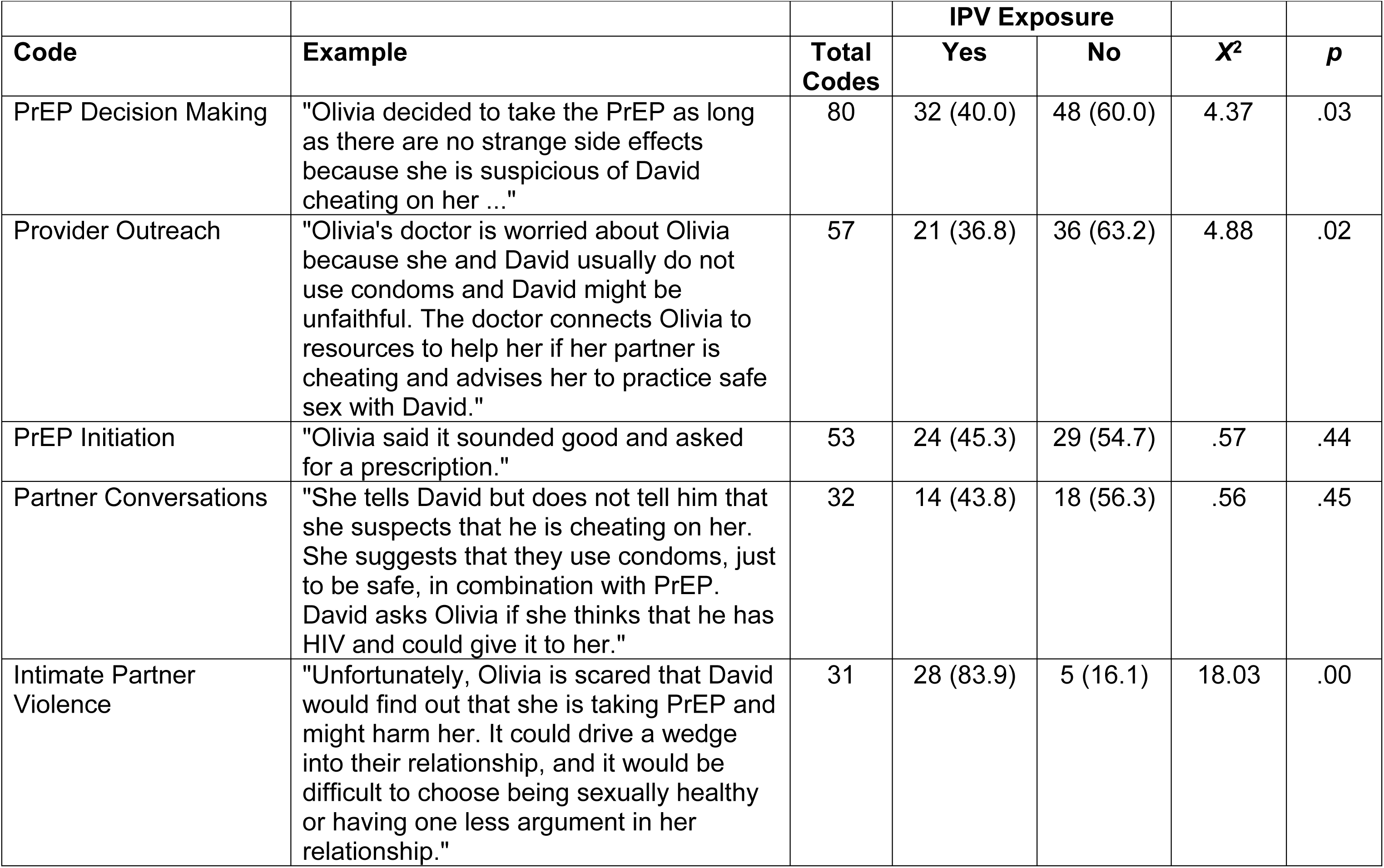
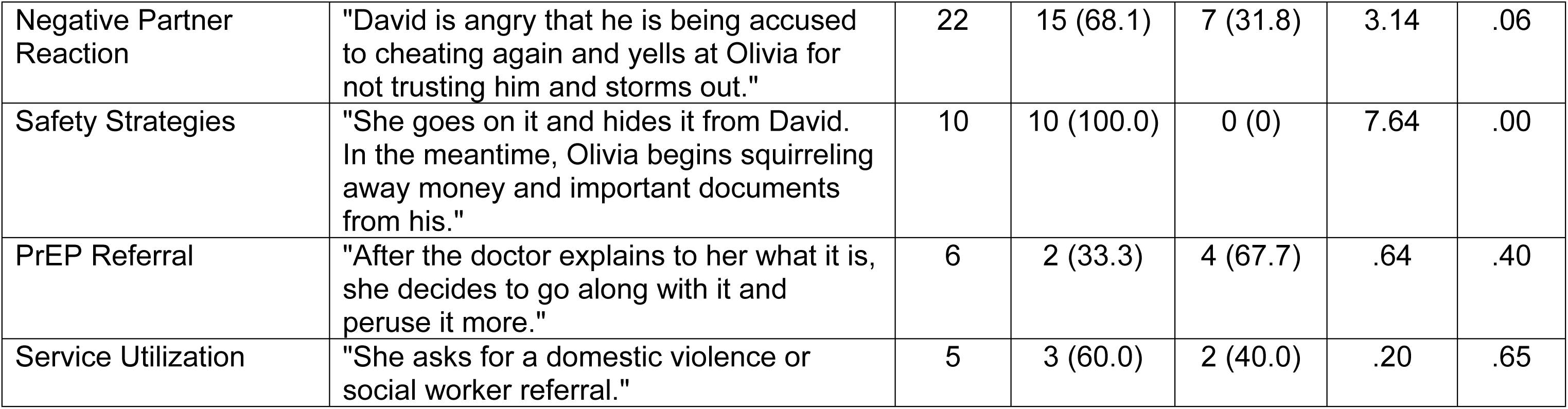
Code Application Frequencies of Nine Most Commonly Used Qualitative Codes among Women With and Without Experiences of Intimate Partner Violence Who Participated in the Prospective Cohort and Story Completion Exercise, United States, 2017-2018.

| Code | Example | Total Codes | IPV Exposure | | $\chi^2$ | *p* |
| --- | --- | --- | --- | --- | --- | --- |
|  |  |  | Yes | No |  |  |
| PrEP Decision Making | "Olivia decided to take the PrEP as long as there are no strange side effects because she is suspicious of David cheating on her ..." | 80 | 32 (40.0) | 48 (60.0) | 4.37 | .03 |
| Provider Outreach | "Olivia's doctor is worried about Olivia because she and David usually do not use condoms and David might be unfaithful. The doctor connects Olivia to resources to help her if her partner is cheating and advises her to practice safe sex with David." | 57 | 21 (36.8) | 36 (63.2) | 4.88 | .02 |
| PrEP Initiation | "Olivia said it sounded good and asked for a prescription." | 53 | 24 (45.3) | 29 (54.7) | .57 | .44 |
| Partner Conversations | "She tells David but does not tell him that she suspects that he is cheating on her. She suggests that they use condoms, just to be safe, in combination with PrEP. David asks Olivia if she thinks that he has HIV and could give it to her." | 32 | 14 (43.8) | 18 (56.3) | .56 | .45 |
| Intimate Partner Violence | "Unfortunately, Olivia is scared that David would find out that she is taking PrEP and might harm her. It could drive a wedge into their relationship, and it would be difficult to choose being sexually healthy or having one less argument in her relationship." | 31 | 28 (83.9) | 5 (16.1) | 18.03 | .00 |
| Negative Partner Reaction | "David is angry that he is being accused to cheating again and yells at Olivia for not trusting him and storms out." | 22 | 15 (68.1) | 7 (31.8) | 3.14 | .06 |
| Safety Strategies | "She goes on it and hides it from David. In the meantime, Olivia begins squirreling away money and important documents from his." | 10 | 10 (100.0) | 0 (0) | 7.64 | .00 |
| PrEP Referral | "After the doctor explains to her what it is, she decides to go along with it and peruse it more." | 6 | 2 (33.3) | 4 (67.7) | .64 | .40 |
| Service Utilization | "She asks for a domestic violence or social worker referral." | 5 | 3 (60.0) | 2 (40.0) | .20 | .65 |

### Findings from Qualitative Story Completions

The participants’ stories of PrEP interest and initiation in heterosexual relationships aligned with four principal narrative types: 1) the *Informed and Empowered PrEP User*, 2) the *Clandestine PrEP User*, 3) the *Hesitant PrEP Contemplator*, and 4) the *Disenfranchised PrEP Non-User*. In this section, we describe each narrative and their plots. The excerpts below demonstrate the different story elements in each narrative.

### Narrative 1: Informed and Empowered PrEP User

Many stories started off with Olivia getting evidence that David was committing infidelity. There were different ways in which Olivia found this information including reading David’s messages on social media and walking in after a long day of work to “catch” him in the act:

> “As she was taking off her shoes, she heard loud thumping in the rooms upstairs. Nervous, she creeps over to the kitchen and unsheathes a kitchen knife from its holder. Quietly, she goes up the steps to investigate the thumping and realizes what she was hearing wasn’t a murderer hiding in her closet. David was cheating on her.” (IPV Experienced Participant, Version B Story)

After finding out that David was committing infidelity, Olivia was often narrated as having an emotional conversation with David about how disappointed she was in his behavior and choices to engage in sexual acts with another female partner. David’s responses are equally narrated as either positive or negative response to this conversation. David’s positive responses are described as reassuring that he is not cheating or getting an STI test as proof:

> “Olivia goes home and tells David he should go to the doctors and get checked out. David is hesitant at first but after getting [information] about PrEP he decides maybe Olivia is onto something and this is good for his health.”(IPV Experienced Participant, Version A Story)

On the other hand, David’s negative responses are described as getting angry, ending the relationship due to mistrust, or becoming physically violent with Olivia: “David immediately questions Olivia when he sees her next and is clearly having trouble keeping a calm composure. David steps to her.”(IPV Experienced Participant, Version B Story) Regardless of David’s response, Olivia remains steadfast on using PrEP to protect her sexual self.

### Narrative 2: Clandestine PrEP User

The most dominant narrative in the stories with Olivia initiating PrEP was the clandestine PrEP user narrative. This narrative describes Olivia getting PrEP and continuing to use it without David learning about her medication. For most of these stories, Olivia’s main conflict is that PrEP is a good option for her due to her concerns about David’s infidelity, but she is also fearful of his reaction to her using this HIV prevention strategy. As a result, Olivia avoids telling David about her PrEP medication

> “She gets the prescription for the HIV prevention pill and begins taking it daily when she takes her birth control. However, she decides not to tell David about it because she is afraid that they will fight, since everytime she asks him about other affairs he gets angry. She is worried that David might find out that she is taking the pill, but she is careful about hiding it from him. As time goes on, she gets more nervous about David finding out that she is on PrEP and is worried about how he might react. She doesn’t want him to get angry or offended, but at the same time she wants to protect herself.”(IPV Experienced Participant, Version B Story)

In some stories, she has developed safety strategies to prevent him from finding out about her medication: “She meets up with David and tells him she’s taking birth control. David doesn’t care because he doesn’t want to get Olivia pregnant.”(IPV Experienced Participant, Version A Story) This narrative generally ends with a positive resolution for Olivia through which she is protected against HIV:

> “While at the physician, she also asks for a referral for a therapist as she would like to work on her self-esteem in order to see if she would like to either work on her relationship with David or find the strength to leave. The physician provides Olivia with her requests and she feels a small sense of security in protecting herself from HIV.”(IPV Experienced Participant, Version B Story)

In contrast, some stories have a climax in which David finds out about her medication towards the end. In this minority of stories, Olivia tells David about her medication directly: “Olivia told David about [PrEP] so that he knows because of his cheating…She boldly confronts him”(IPV Experienced Participant, Version B Story) or David finds out by watching her behavior change: “Days later he sees her taking the pills. He becomes suspicious. Now David is snooping. And Olivia is hiding.”(No IPV Experience Participant, Version A Story) These stories tended to end with David becoming angry or physically violent with Olivia. Similarly, this storyline equally ends with either a positive or negative resolution for Olivia. In the positive resolutions, Olivia ends the relationship with David, lives a healthy life, and finds a new loving partner or travels the world: “They break up. She decide she wants to travel. Visit the world. She finds a new career and lives happy ever after”(No IPV Experience Participant, Version A Story). However, in the negative resolutions, David’s violent behavior become life-threatening for Olivia: “He got mad that Olivia got the drug. So he confronted her and beat her so bad that Olivia was bleeding everywhere. Olivia managed to get away and call 911.”(IPV Experienced Participant, Version B Story)

### Narrative 3: Hesitant PrEP Contemplator

The third narrative was less prevalent but still captured several participants’ stories: the hesitant PrEP non-user narrative. This narrative often described Olivia as moving between PrEP pre-contemplation to the PrEP contemplation stages in her decision-making process. The main conflict in this narrative was the fact that Olivia did not want to cause any additional relationship problems with David and thought that PrEP would agitate their relationship dynamic.

In this narrative, Olivia is narrated as learning about PrEP from her doctor and needs more time to process whether this a good option for her: “Olivia finds out more about the treatment because it is a noninvasive drug that can help prevent against HIV. She tries to find out more about this drug…”(IPV Experienced Participant, Version B Story). After leaving the doctor, Olivia is deciding whether to discuss her new information about PrEP with David. In most of these stories, Olivia does not discuss PrEP with David because she is fearful of his reaction:

> “She returns home and wonders if she should discuss what happened at the doctors today with David but decides she should probably keep it to herself because it might make David upset. Olivia wants to think it over for a few more days and see where her head is at with all of this.”(No IPV Experience Participant, Version A Story)

This narrative usually ends with an ambiguous resolution such that it is unclear if Olivia ever uses PrEP or some type of HIV prevention strategy.

In contrast, some stories have a climax in which Olivia discusses her new information about PrEP with David and even asks to use alternative HIV prevention strategies such as condoms: “Olivia decides to have an open conversation with David. After their conversation Olivia decided to begin using condoms and may consider using the HIV prevention drug because you can never be to careful.”(No IPV Experience Participant, Version A Story) In this alternate storyline, David generally responds negatively:

> “David did not want to do this so he threatened to leave Olivia if she made him use a condom. Olivia told him that if he cared for her at all, that her request would not be a big deal. She told him that she loved him, but she loved herself more. Olivia decided that she would allow David to go and just be single. About 6 months later, Olivia spoke with a relative of David’s, who told her that he was diagnosed with HIV and was depressed. She was sad to hear that, but was happy that she chose herself.”(No IPV Experience Participant, Version A Story)

### Narrative 4: Disenfranchised PrEP Non-User

The final narrative, the disenfranchised PrEP non-user, characterizes Olivia has remaining a PrEP non-user, often due to external forces beyond her control. In this narrative, Olivia learns about PrEP during her doctor’s visit; however, there are clinical and social barriers she experiences during her PrEP decision-making. Specifically, Olivia does not like the side effects associated with PrEP: “Oliva inquiries about the drug but decides not to take it because she does not have HIV and learns about the frightening side effects”(No IPV Experience Participant, Version A Story); or has unfavorable opinions of pharmaceutical companies:

> “Olivia asks her doctor what PrEP is, what it is used for, and what the side effects are. She then does her own research because pharmaceutical companies have become too commercialized and even doctors try to sell you things.”(No IPV Experience Participant, Version A Story)

In addition to clinical barriers, in some stories, Olivia faces social barriers such as extreme fear of David’s reaction:

> “Olivia then proceeds to learn more information from her doctor about this new drug. She is afraid to tell David about all of this. Unfortunately, Olivia puts it all out of her mind in fear that it’s not worth it.”(IPV Experienced Participant, Version B Story)

Similarly, Olivia also has low perceptions of her PrEP candidacy because she believes PrEP is for different key priority populations: “Olivia still feels uncomfortable with the idea of using this because she feels it is something for prostitutes or intravenous drug users, not people suspecting an unfaithful partner.”(No IPV Experience Participant, Version A Story) Overall, these clinical and social barriers are the main conflicts for Olivia in this narrative. The storyline usually lacks a climax because she avoids confronting these barriers. This storyline also is characterized with an ambiguous resolution for Olivia because there’s no definitive conclusion on her use of HIV prevention strategies.

In contrast, a few stories within this narrative includes David finding out about her clinical discussion regarding PrEP either by seeing patient education materials or Olivia discussing the topic directly with David:

> “Olivia takes home the pamphlet to look over and decide if she thinks that it is right for her. She puts it in the nightstand drawer. David finds the pamphlet the next day while looking for something in Olivia’s drawer. He becomes enraged asking Olivia what she needs this drug for, and has she been cheating on him. She tries to explain it’s just precautionary, just in case. David then gets more upset, saying she’s basically accusing him of cheating on her. She begins to ask him if he has. He denies it and says awful things like how just because Olivia is a slut, whore etc doesn’t mean he goes out cheating. He eventually becomes so angry he begins hitting Olivia over it. This makes Olivia more unsure if she should take the drug as a precaution or avoid it so that David doesn’t have a “reason” to be aggressive towards her.”(IPV Experienced Participant, Version B Story)

In each of these alternative stories, Olivia and David have an argument and dissolve their relationship. The resolutions have been both positive and negative for Olivia in these alternate stories, however, the negative resolution is more dominant. An example of a positive resolution includes she is tested negative for HIV and will use HIV prevention methods in the future:

> “Olivia thinks about what her doctor told her and realized that her and her boyfriend will start using condoms. She meets up with her boyfriend and tells him she wants to [use] condoms; he becomes angry and puts his hands on her again. Olivia starts to cry, [and] David tells her he’s sorry. She sits still and quietly for ten minutes, starts to boil hot water. David pleading in the background saying sorry and he won’t do it again. She throws the hot water on him and tell him to leave the apartment and never come back again. Few days later she gets her test result back and everything was negative. She moved on with her life but she decided to use condoms more because she doesn’t want to risk her life anymore.”(IPV Experienced Participant, Version B Story)

However, a negative resolution includes Olivia being physically abused by David and also Olivia being tested positive for HIV: “But in the end once they break up she wants to be on the pill but it is too late because she caught an unpleasant disease [HIV] that she regrets for the rest of her life.”(IPV Experienced Participant, Version B Story)

## DISCUSSION

Reducing the PrEP effectiveness-implementation gap among U.S. cisgender adult women is a critical public health priority.^11,14,39^ To date, only 10% of cisgender women who could benefit from PrEP are actually prescribed this novel HIV prevention medication.^40^ While several studies have documented the structural barriers to PrEP access among women,^4,15,16^ our study is the first to leverage story completions, a novel qualitative methodology rooted in psychoanalytic projective techniques,^29^ which aims to gain psychological and sociocultural meaning to PrEP interest and initiation in heterosexually-active relationships among cisgender women. In our sample, we identified four main narratives that addressed the potential complexities of accessing, advocating, and initiating PrEP as a cisgender woman in a heterosexually-active relationship. Several of the narratives align with extant research demonstrating that fear of male partner’s reaction, distrust in the medical system, secrecy, and low HIV risk perception contribute to little or no interest in PrEP.^2,10,41–43^ However, the narratives that demonstrated women’s empowerment and agency offers a promising avenue for future PrEP-related communications and interventions for cisgender women in heterosexually-active relationships.

Consistent with previous research on women’s empowerment and agency in PrEP initiation,^44–46^ the *Informed and Empowered PrEP User* narrative characterized Olivia as the heroine in her own story who centered her sexual needs regardless of her male partner’s reactions. Extant research with PrEP-naïve women indicate that being concerned about a negative reaction from a male partner, especially among women experiencing IPV, was a key interpersonal barrier to PrEP interest and initiation.^10,47,48^ Moreover, qualitative studies with PrEP-engaged women often discuss this population as being resilient and prioritizing control of their sexual health as key facilitators to initiation.^44,45^ Altogether, this narrative reflects a prominent conceptualization of PrEP interest and initiation in which women actively resist the socialized power inequity between women and men at the interpersonal level in order to be empowered to pursue their desired direction and vision. While prior research indicates that marketing can influence women’s perceptions of appropriate PrEP candidates,^2,49,50^ our findings support the potential of developing empowerment-based PrEP interventions and reframing PrEP-related communications to women’s empowerment and agency for cisgender women.

A dominant narrative was the *Clandestine PrEP User,* which describes our main female character as exhibiting agency but also utilizing survival and protective mechanisms to use PrEP safely. As a result, she uses different tactics to conceal her use of PrEP to reduce the likelihood of relationship dissolution and violent retaliation from her partner. This narrative aligns with prior research suggesting that PrEP-engaged women may hide their oral PrEP pills or pretend the medicine are vitamins to reduce anticipated stigmatizing and negative reactions from partner, family, and friends.^44,51,52^ Similarly, a participatory action research study among women involved in a PrEP clinical trial found that maintaining product secrecy was a key theme for product adoption.^41^ Coupled with previous research, our findings suggest that women may view the concept of PrEP in heterosexually-active relationships as personally beneficial, but socially stigmatizing and threatening to their partners. Thus, to protect themselves and maintain their intimate relationship, women that perceive protective mechanisms while using PrEP as the best route. While women’s empowerment is stifled in this narrative, it may be useful for healthcare practitioners to acknowledge this complicated dynamic for cisgender women in heterosexually-active relationships, especially women experiencing IPV, and discuss engaging in protective mechanisms while using PrEP.

The last two narratives, the *Hesitant PrEP Contemplator* and the *Disenfranchised PrEP Non-User*, described our main female character as not using PrEP for two main reasons: contemplation and sociostructural factors. Prior studies indicate that sociostructural factors such as distrust in pharmaceutical companies and non-inclusive PrEP marketing have contributed to a lack of PrEP interest among cisgender women.^2^ Furthermore, earlier qualitative studies found that women needed additional information to make a decision regarding PrEP^49^ and potentially decision aids to help support this process.^53^ Overall, a greater investment in preventive interventions is needed to address and change these current conceptualizations of PrEP in heterosexually-active relationships. For example, more opportunities to increase PrEP education and decision tools in settings where cisgender women in heterosexually-active relationships frequent, develop inclusive marketing, and incorporate gender-transformative programming that redress the power inequality between women and men.

Our study also found that some codes were applied differentially among stories written by women with and without IPV experiences. In particular, PrEP decision-making and Provider outreach were applied in stories written by women without IPV experiences, whereas IPV and Safety strategies were applied in stories written by women with IPV experiences. Since coders were unaware of participant’s characteristics and story stem version, and bias was reduced with randomization, it is possible that these differential applications of qualitative codes reflect the actual representations of PrEP from women’s perspectives. Specifically, women without IPV experiences may conceptualize situations in which women are making personal decisions about PrEP likely with the help of a healthcare provider. On the other hand, women with IPV experiences are viewing IPV perpetration as an obstacle to PrEP initiation and the need to have safety strategies in place for women who decide to use PrEP. Trauma-informed practices should be coordinated with PrEP-related communications, campaigns, and interventions to increase women’s PrEP adoption.^11^

The study’s findings should be interpreted considering the following limitations. Our story completions exercise was based on a hypothetical situation written in third person to illicit women’s conceptualizations and understandings. While it is possible that our participants had personal experiences of sexual infidelity, the writing exercise was not about their actual lived experience and most of our sample were not prescribed PrEP at the time of the follow-up data collection. IPV was self-reported and can be influenced by social desirability bias, such that the prevalence of IPV could be under-reported. While self-report measures for IPV are the most common methods,^54^ this study only accounted for the exposure to IPV, not the frequency and severity. The current study was conducted five years after the FDA approval of daily oral PrEP, and thus the study was focused on this PrEP modality. However, the approval of long-acting injectable at the end of 2021 may address some clinical barriers to PrEP initiation for cisgender women. Our study was conducted with cisgender women in Connecticut. To increase transferability of the qualitative findings to other similar contexts,^55^ we provided rich descriptions of the original research to aid other researchers to determine whether these findings will be unique to their setting.

## CONCLUSIONS

This study provides evidence on the different representations and understandings of PrEP interest and initiation for cisgender women in heterosexually-active relationships. Our four narratives show how complex women view their ability to access and use PrEP, while navigating the anticipated reactions from the interpersonal actors (e.g., male partners) and social institutions (e.g., pharmaceutical companies). If our findings are replicated in studies in different settings, it will provide substantial support for future prevention interventions adopting empowerment-centered approaches to refocus women’s needs in the context of PrEP initiation and healthy relationships.

## Data Availability

Data cannot be shared publicly because of participants' privacy.

